# The Response of Pakistani Social Workers amid the COVID-19 Pandemic: A Qualitative Analysis of the Main Challenges

**DOI:** 10.1101/2020.10.18.20214601

**Authors:** Kiran Abbas, Muhammad Inam Ul Haq, Wareesha Afaq Zaidi, Ahmed Kaleem, Hamza Sohail, Moiz Ahmed

## Abstract

The study aimed to highlight the main challenges faced by the social workers amid the pandemic. A qualitative study was conducted between March 2020 to May 2020 in Karachi, Pakistan. All participants who belonged to a non-profit organization were eligible to participate. Open-ended questions were asked by the participants. The mean age of the participants was 24.8 ± 5.9 years. The main challenges faced by the social workers were: i) resistance from the family and friends, ii) lack of personal protective equipment, iii) mistrust from public, iv) uncooperative government/authorities.

## Introduction

As the Covid-19 pandemic rages on, the burden on social workers has immensely increased.^1^ The most prominent of these responsibilities include caring for the diagnosed Covid-19 patients in isolation camps, providing them food and water, making burial arrangements for the patients who died of COVID-19, and providing relief health and food packages to the residents of poor socioeconomic localities in Pakistan.^2^

Not only the social workers are at high-risk of contracting the virus themselves, but their families are also exposed.^2-3^ These unsung heroes are also at risk of mental health issues due to stress, anxiety, and work-related exhaustion.^4-5^ The situation is worse in developing countries including Pakistan, India, Nepal, among others. These countries do not have enough resources to take care of the rapidly increasing demands as more cases of Covid-19 are reported. The health-care sector in Pakistan, is already supersaturated lacking sufficient “resources” including the machines, drugs, facilities, supplies and workforce.^5^ Many hospitals refuse to admit patients of Covid-19 as they are not equipped to handle them. There is a shortage of personal protective equipment in many hospitals and doctors are rendered exposed to this dangerous virus.^6^ Many households are now facing food shortages as the majority of the Pakistani people are daily wage earners. Due to the lockdown, they are facing unemployment for the past sixty to ninety days.^7^ This is where social workers play a huge and impeccable role in our society. From collecting the donations from the privileged sector of the community, to bulk-buying personal protective equipments, groceries, and clothes, to distributing them to the health-care workers who are battling the Covid-19 pandemic at the frontlines and ensuring adequate distribution of groceries (rations) and other essentials to families in need, social workers are continuously and relentlessly serving our global communities.^5-8^

Social work is as equally important as health-care work, and both work in synergy. Nevertheless, many studies have highlighted the impact of pandemic solely on the health-care practitioners^2-4^, however, the problems faced by the social workers remained unexplored till now. In this brief study, we highlighted the main issues concerning the social workers and how they are dealt with in Pakistan amid the Covid-19 pandemic.

## Methods and Materials

A qualitative study was conducted between March 2020 to May 2020 in Karachi, Pakistan. After obtaining clearance from the institutional ethics review committee, the participants were enrolled using non-probability, convenience sampling. A total of twenty participants were interviewed using call-based short surveys. All participants who belonged to a non-profit organization falling within the definition of social work as previously mentioned were eligible to participate. All participants not willing to participate in the call-based interviews were excluded from the study.

Qualitative data was based on a brief interview which took place over a phone call or video call as per feasibility of the participant. The call was recorded after taking verbal and electronic consent from the participants. The interview was focused on evaluating the main challenges faced by the social workers amid the COVID-19 pandemic and they dealt effectively with those challenges. Open-ended questions were asked. The entire discussion was recorded and then transcribed verbatim. The discussion was conducted in English and transcripts were produced in English language.

### Statistical Analysis

The age of the participant, length of the interview, and the work hours were presented as mean plus standard deviation while gender was presented as frequency/percentage.

Microsoft Excel sheet was used for data analysis. Qualitative data was analyzed using an inductive approach. The transcribed data was organised, main themes were identified, data was labelled and finally coding was done. The codes and themes were then analysed and presented.

### Ethics

All participants gave informed verbal and electronic consent prior to the interview. Participants were ensured that complete anonymity and confidentiality would be maintained. The participants were given pseudonyms for transcription purposes to ensure their anonymity. Apart from the principal investigator Abbas K, no one had access to the original interviews.

## Results and Discussion

We interviewed twenty social workers randomly selected from different organizations currently, active and functional. The mean ± SD interview time was 21.4 ± 10.5 minutes. There were 6 (30%) female while 14 (70%) were male social workers. The mean age of the participants was 24.8 ± 5.9 years. The main themes explored are listed in Table 1.

### Psychological Impact on Social Workers amid COVID-19 pandemic

Apart from the challenges faced by the social workers, many experienced psychological effects as a result of the pandemic. The mean hours the participants worked were 7.9 ± 5.6 hours per day. This amounted to approximately forty hours a week excluding the weekends. The participants claimed that they were constantly terrified of contracting the virus themselves and also the fear of infecting their family members takes a toll on their mental health. 19 out of 20 (95%) participants claimed that they were more stressed out than usual, 8 (40%) self-reported insomnia and sleep disturbances, while 14 (70%) claimed they were fatigued after working long hours.

### Coping mechanism to deal with Challenges

The majority of the participants who were interviewed agreed that in spite of the apparent challenges faced by social workers, they would continue to help others. The one single-most important driving force was the thought of helping the less-privileged community during the pandemic. They realized that many daily wage earners were out of employment hence,they had no means to put bread on the table. Therefore, social workers must play their parts in helping out the community in these awful times. One participant claimed, “The fact that I am able to help others by providing them with groceries and clothes is enough for me - I do not expect any reward for my social work.” Another social worker claimed that, “I am not wasting my time binge-watching TV series on netflix like others but actually doing something useful. It gives me motivation.”

As it was previously pointed out that the long work hours lead to fatigue and burn-out among social workers therefore, many recommended taking mini-breaks in between work hours to prevent burn out. One young social worker claimed, “We are prone to fatigue as we are fasting during the holy month of Ramadan - therefore we take short breaks of 10-15 minutes to prevent full-fledged burn-out - it helps keep us productive.”

Another main concern reported by the social workers was the fear of infecting the family members. Therefore, it was found that 12 (60%) social workers were not directly in contact with their family members. They were restricted to their own rooms - to keeping their family members safe from the viral pandemic.

### Challenges faced by Social Workers amid COVID-19 Pandemic

The first theme was “Challenges faced by Social Workers”. As many as 13 out of 20 participants divulged that their family members were not in support of them going out in the field and exposing themselves and their families to the virus. The family members were concerned about the safety of their loved ones. However, some participants also remarked that despite the fear and associated anxiety, their family members fully supported their cause and encouraged them. One social worker said, “I believe that without the support and encouragement of my family, I would not be able to perform my duties. I owe them so much.” Another participant claimed, “My family was terrified in the beginning - they made me wear all sorts of protective equipment before I could go out in the field, but with time they realized that the work I did was so important - I was helping other families during these unpredictable awful times. With time, they started supporting me and I am grateful for that.”

The second challenging aspect that was faced by the social workers was the non-availability of personal protective equipment (PPE). Over 10 (50%), participants claimed that they do not wear any personal protective equipment during work hours. The main reasons divulged by the participants were: shortage of PPEs and unnecessary financial burden on organization. The interviewee explained that since the inception of lockdown, prices of these essential PPEs have drastically increased. Hence, the small student-based organizations were unable to provide their workforce with basic PPE. 14 participants used only surgical masks, 3 participants used both surgical masks and gloves, while two participants worked without any PPE in the field. Upon asking one social worker said, “We tried to follow the standard operating procedures as laid by the Government of Pakistan but eventually we were unable to sustain the availability of PPE for our volunteers. However, even then these remarkable people volunteered and risked their lives for the sake of humanity.”

Another social worker claimed, “We take all necessary precautions - wash our hands, use sanitizers, avoid contact as much as possible, follow respiratory hygiene among other preventive measures - but we also know that if we do not go out to supply food bags to these poor people, they may die of hunger or commit a crime.”

Many social workers were young and inexperienced, but their spirit was commendable. They were all highly motivated to help their fellow citizens. The female social workers claimed that their main role was to do public marketing and were responsible for the packaging of the donations (PPE, food/ration bags/clothes, etc.). The main challenge for these young social workers was the mistrust of the public. Since the start of the pandemic, the participants revealed that some fraudulent illegitimate organizations have surfaced which collect money from the public in the name of helping the less unfortunate and then disappear. One person claimed, “At the start of our donation drive, many people did not trust us. It felt bad. So, we used social media i.e. Instagram, facebook, Whatsapp, among other softwares to spread the word across the nation. People started appreciating the work we did and the rate of donations increased tremendously ever since.”

Incorporating the use of media to build-up trust among the general public was one of the strategies employed by the social workers to ensure steady supply of donations. Another strategy was to offer a pick-up service to the person who was interested in donating in cash. Since, many people were scared of being scammed online.

None of the social workers faced shortage of manpower. There were enough willing participants and volunteers who also shared the same vision as the organization they were associated with i.e. to help the deserving people of the country. One person said, “We always had enough people on board to help others, we were never short of volunteers.”

Another main hindrance faced during the social work was differentiating the families who were really deserving and those who were not. “Many times, we were contacted by elite/well-off families to donate them groceries at their doorsteps, it was disgusting to see this kind of behavior from people.” It was necessary to verify whether the person or family was actually underprivileged or not. For that, many non-profit organizations did a background search or asked for some justification.

The social workers were discouraged by the non-cooperative stance of the government and politicians. One social worker reported, “We tried to contact one MNA, to ask for donation but he did not pick-up his call.” (MNA is short for ‘Member of the National Assembly)

Not many social workers were concerned about the security. Since the female workers did not participate in the field work, it was not a big challenge. However, the social workers claimed that the Pakistan Rangers (Sindh) provided them security upon request. Pakistan Rangers are a respectable force which has earned a great repute among people.

The ministry of health and services, Pakistan has published guidelines on the proper use of disposable face masks/surgical face mask/medical face mask which is one of the basic PPE. It is often used by the patient in healthcare settings to protect others from a communicable infection. New developments regarding COVID-19 have shown that a patient may be infected with COVID-19 virus and still appear well or asymptomatic for a certain period before experiencing the symptoms. Meanwhile, is completely capable of shedding virus during sneezing, coughing and while speaking. Therefore, it is prudent for the public to wear face masks with an intention to protect others and themselves from droplets of saliva/sputum particularly when entering closed or congested areas. Many countries have adopted this as a national guideline.^9-11^

In our study, the social workers explained that the prices of even the basic PPE including surgical masks or gloves have accelerated by many times hence, it became impossible for small student-based organizations to keep supplying their volunteers with these facilities.^12^ According to a CNN report, the cost of PPE is continuously skyrocketing, in some cases by over a 1000 percent.^13^ Some social workers, mainly focused on distributing PPEs to health-care practitioners including the N95 respirator, gowns, gloves, medical mask, and eye protection equipment.^14-15^

The main objective of these organizations was to ensure that the poor and the underprivileged sector of the community were provided with groceries and other essential items. Hence, these organizations targeted the low-socioeconomic areas including the shanty towns, villages, and nursing homes all across the country.

This is the first report which highlighted the new emerging challenges during COVID-19 pandemic faced by the social workers in Pakistan. The government and concerned authorities must cooperate with these social workers so that they can help their country in the times of need.

## Conclusion

The majority of the social workers were young and inexperienced, but their spirit was commendable. They were all highly motivated to help their fellow citizens. We found that with the help of the government and health authorities, they could achieve more success in helping their fellow citizens.

## Data Availability

The data for the study includes personal identifiers and no one has access to the data apart from the principal author. Decoded data can be provided if requested. Kindly contact principal author through

## Acknowledgements (if any)

We would like to acknowledge the social workers in Pakistan who participated in this study and are serving the country amid the crisis.

